# Comparing Single-Cell Transcriptomes of Blood and Cerebrospinal Fluid Leukocytes in Multiple Sclerosis

**DOI:** 10.1101/2024.05.09.24307127

**Authors:** Saed Sayad, Mark Hiatt, Hazem Mustafa

## Abstract

**Background:** Multiple sclerosis (MS) is a chronic autoimmune disorder of the central nervous system, marked by inflammation, demyelination, and neurodegeneration. Diagnosis is complex due to overlapping symptoms with other neurological conditions, typically relying on clinical evaluation, neurological exams, and tests like magnetic resonance imaging (MRI) and cerebrospinal fluid (CSF) analysis. Recent advances in technology, particularly single-cell analysis of blood and CSF leukocytes, hold promise for enhancing MS diagnosis by providing insights into immune cell involvement at a molecular level, potentially enabling more precise diagnostics and personalized treatments.

**Method:** We acquired single-cell RNA Sequence (RNA-Seq) data (*<u>GSE138266</u>*) from the website of the National Institutes of Health of the United States (NIH), comprising blood and CSF samples from patients diagnosed with idiopathic intracranial hypertension (IIH) and MS. Our analysis focused on identifying genes, pathways and gene ontology terms with distinct expression patterns in MS compared to IIH.

**Results:** We identified clear differences in gene expression profiles between blood and CSF samples in MS, contrasting with single-cell leukocyte samples from IIH. The increased expression of genes in MS suggests a boost in immune activity and regulation of cellular proliferation, while decreased expression points to disruptions across various functional categories. Gene ontology analysis identifies upregulated terms associated with cellular differentiation, apoptotic regulation, and immune responses in MS, while downregulated terms suggest disruptions in cellular signaling cascades and myelination processes. Similarly, Reactome pathway analysis unveils upregulated pathways in MS related to cell cycle regulation and immune mechanisms, contrasting with downregulated pathways indicative of disruptions in oxygen transport and cellular metabolism.

**Conclusion:** Our study offers a thorough examination of single-cell transcriptomic data, unveiling unique gene expression patterns, gene ontology terms, and Reactome pathways linked to MS pathophysiology. Notably, our findings identify *CD69* and *HNRNPK* as potential key genes driving MS progression. By clarifying molecular differences between MS and IIH, our findings enhances our grasp of MS pathogenesis and unveils promising targets for diagnostic and therapeutic interventions.

## Introduction

MS is a chronic autoimmune disorder characterized by inflammation, demyelination (damage to the protective covering of nerve fibers), and neurodegeneration in the central nervous system (CNS), which includes the brain and spinal cord. This complex condition can lead to a wide range of symptoms, including fatigue, weakness, difficulty walking, numbness or tingling, vision problems, pain, and cognitive impairment. The exact cause of MS is still unknown, but it is believed to involve a combination of genetic, environmental, and immune system factors. Diagnosing MS can be challenging due to its diverse and unpredictable symptoms, which can mimic those of other neurological disorders. Medical professionals typically rely on a combination of clinical evaluation, medical history, neurological examination, and diagnostic tests to confirm a diagnosis of MS. These tests may include magnetic resonance imaging (MRI) to detect areas of demyelination and inflammation in the CNS, CSF analysis to check for abnormalities such as elevated levels of certain proteins and immune cells, and evoked potential tests to assess the function of nerve pathways. In recent years, advances in technology have led to the development of new diagnostic tools and techniques for MS, including single-cell analysis of blood and CSF leukocytes [1]. This approach allows researchers to study the immune cells involved in the pathogenesis of MS at a molecular level, providing insights into disease mechanisms and potential biomarkers for diagnosis and prognosis.

## Data Analysis

We obtained single-cell RNA-seq data (*GSE138266*) from the NIH portal website comprising blood and CSF samples from patients diagnosed with IIH and MS (**Figures 1 and 2**).

**Figure 1:**
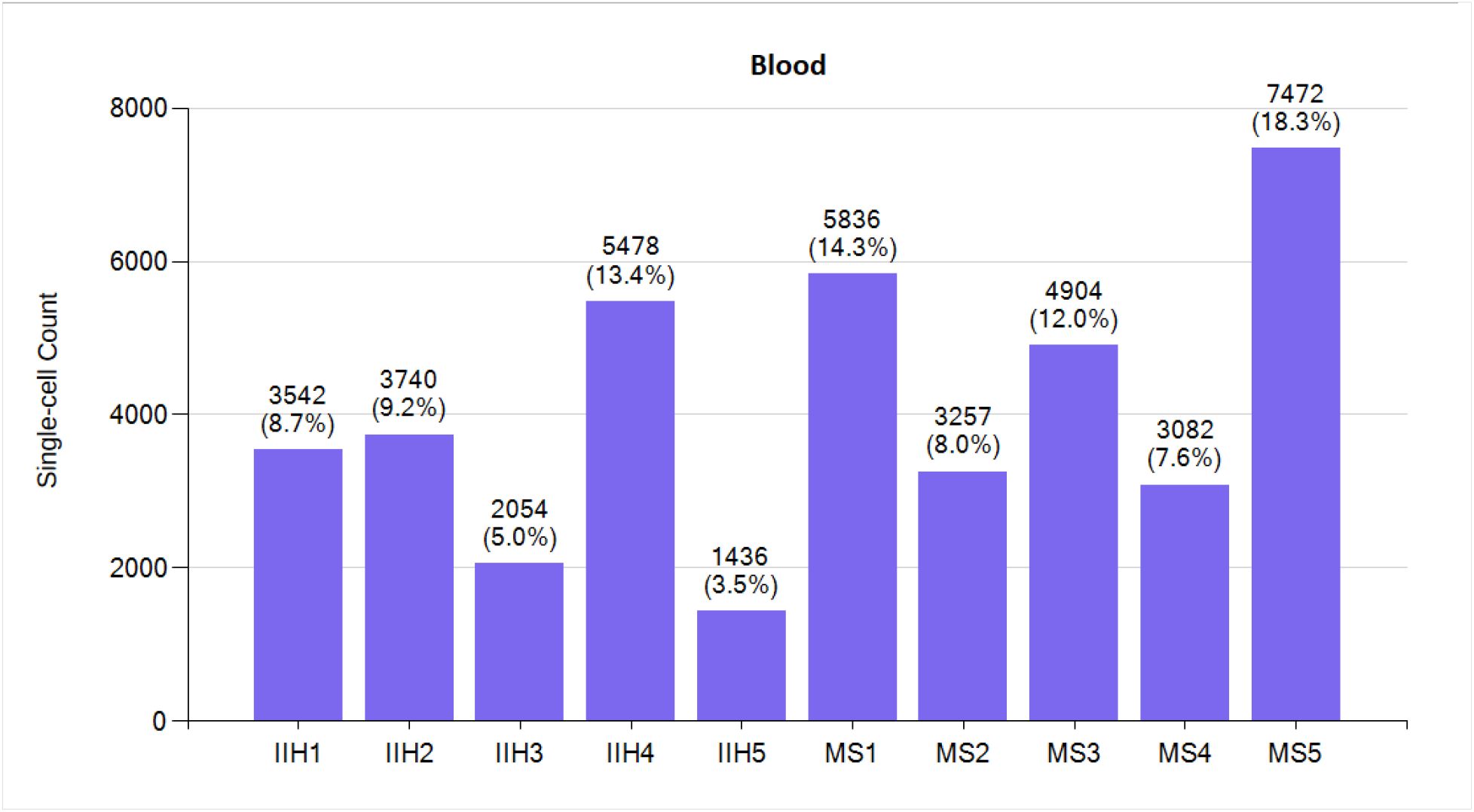
Blood leukocytes single-cell count for five IIH patients and five MS patients.

**Figure 2:**
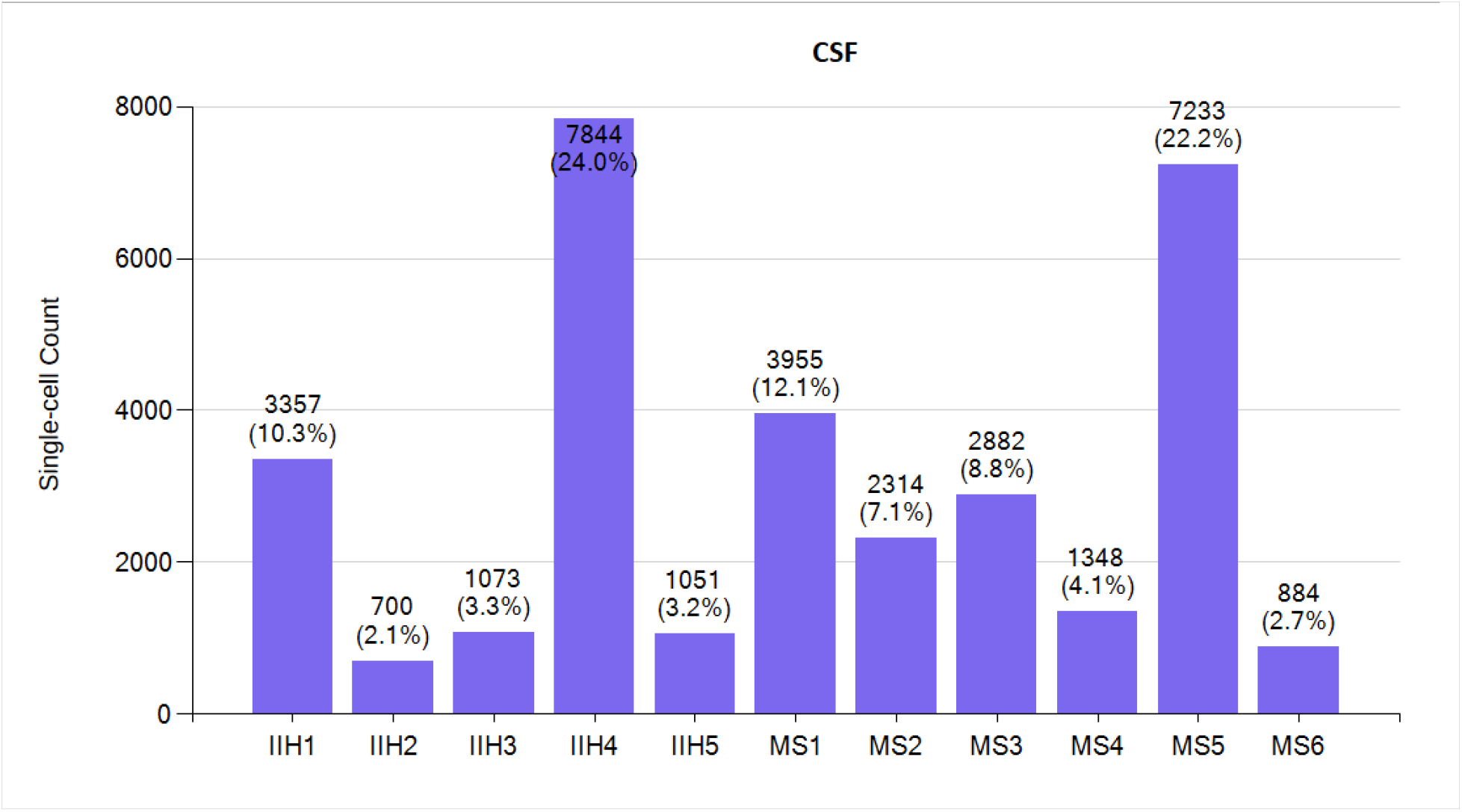
CSF leukocytes single-cell count for five IIH patients and six MS patients.

We excluded data from one patient (GSM4104128) due to an unusually high number of 737,279 barcodes, as it could potentially introduce bias or artifacts into our analysis.

In this study, we have done an extensive comparative analysis focusing on the single-cell transcriptomes of leukocytes isolated from both the peripheral blood and CSF of individuals afflicted with MS and IIH. Our primary objective was to find the gene expression signatures underpinning the pathophysiology of MS. By harnessing the power of single-cell transcriptomes, we were able to interrogate individual cells, thereby uncovering subtle variations in gene expression, pathways and gene ontology profiles. This integrated approach allowed us to have deeper insights into the molecular signatures distinguishing MS from IIH thereby advancing our understanding of MS pathogenesis and potentially paving the way for more precise diagnostic and therapeutic strategies.

### Differential Gene Expression – Comparing Single Cells within a Single Patient

We randomly divided the single-cell data into two subsets for each of the 21 patients, then employed a t-test to identify genes with differential expression. Our thorough examination showed no statistically significant upregulated or downregulated genes within either the IIH or MS patients across blood or CSF samples. This outcome bolsters the reliability of the single-cell RNA-Seq data. Here, we just present the results for one MS patient (MS1) as illustrated in **Figures 3** and **4**.

**Figure 3:**
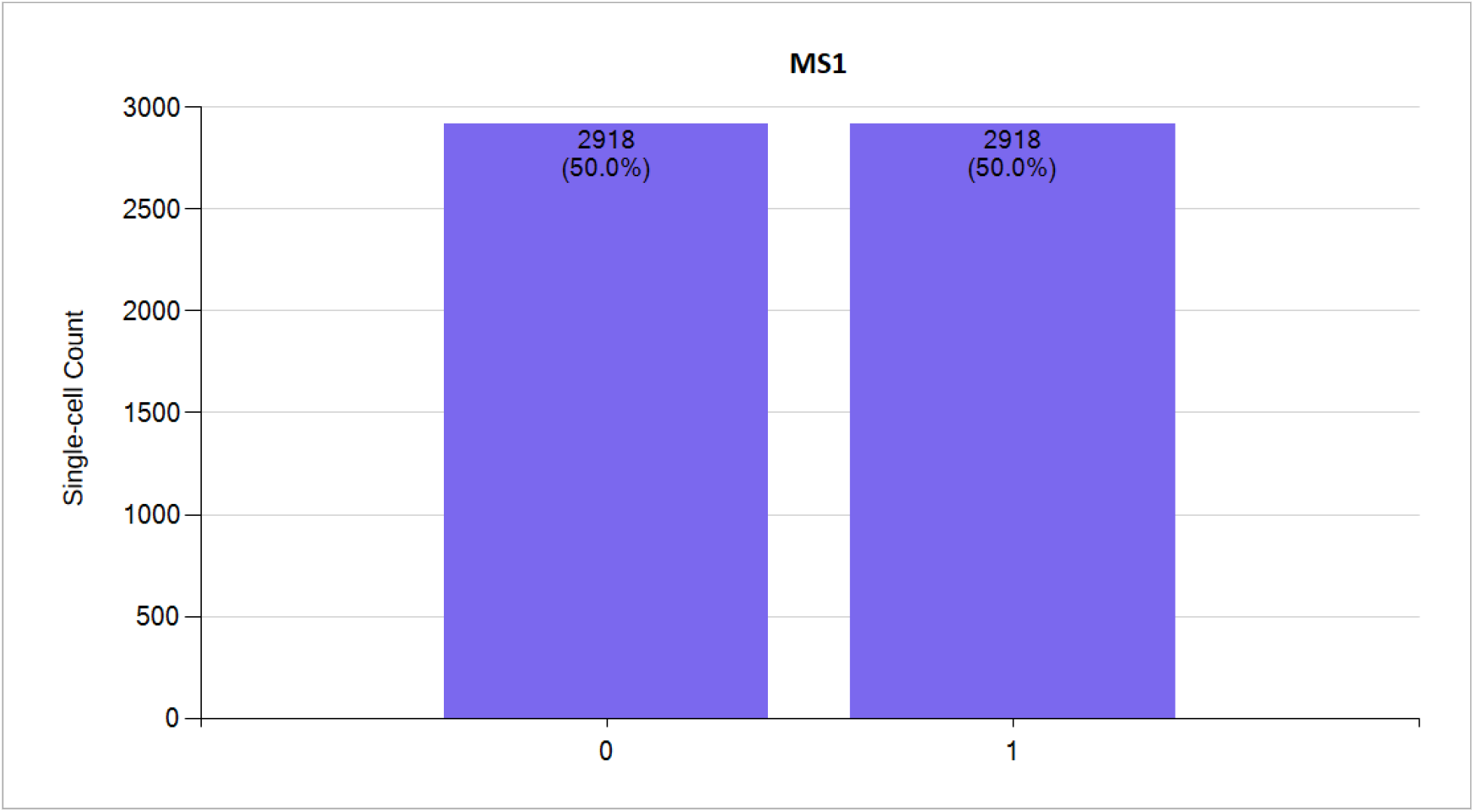
Blood leukocytes single-cell count for MS patient (MS1) divided to two subsets randomly (0,1).

**Figure 4:**
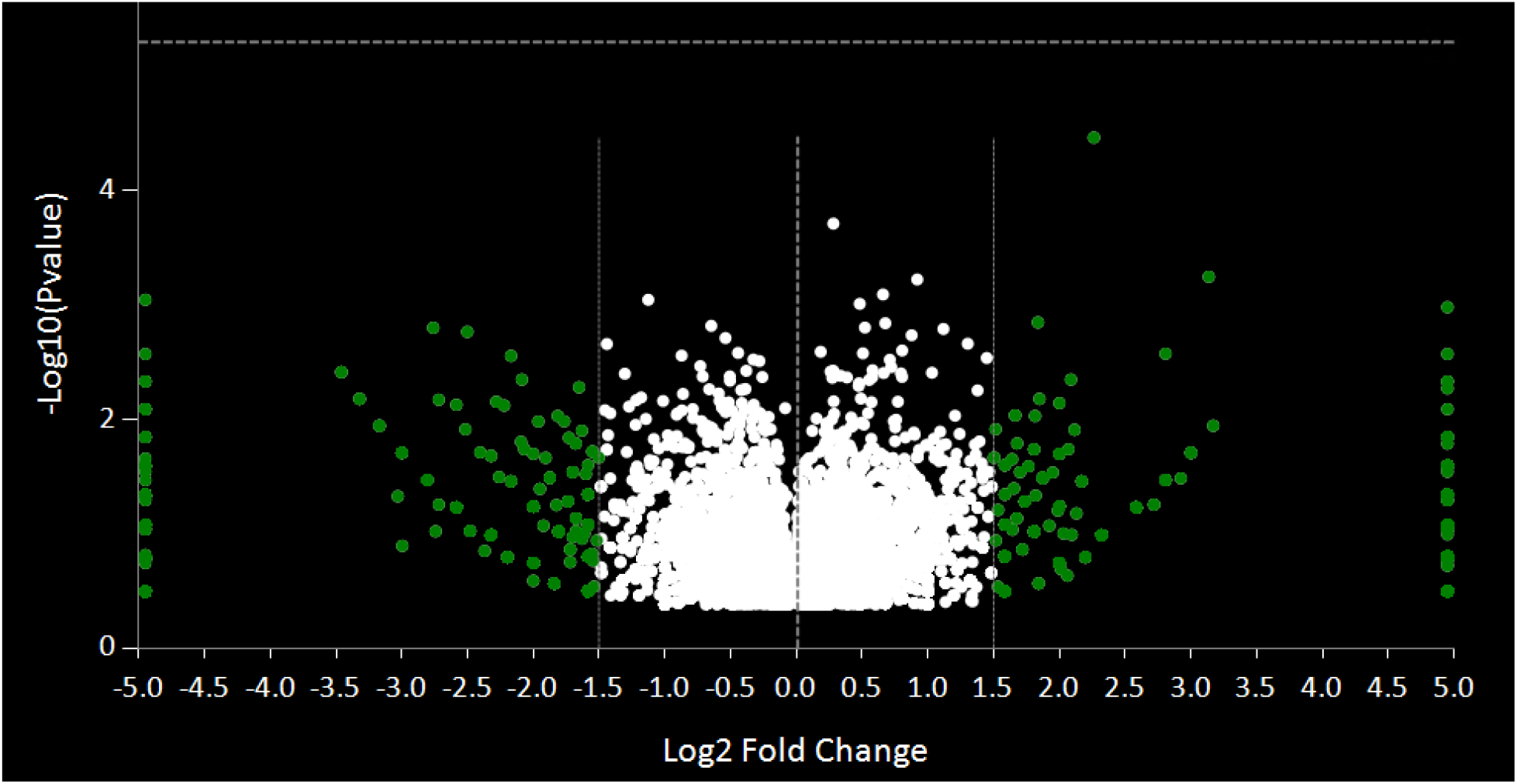
No significant upregulated or downregulated genes were detected between two subsets of blood leukocytes of MS1 patient.

### Differential Gene Expression – Comparing Single Cells within the same Group of Patients

We randomly divided the IIH patients into two subsets and observed no significant upregulation or downregulation of genes in either the blood or CSF groups using t-test. This pattern persisted when we repeated the analysis for all MS patients, where we similarly found no significant differences in gene expression between the subsets. These results underscore the robustness of the single-cell RNA-Seq data, as illustrated in **Figures 5** and **6** for IIH patients.

**Figure 5:**
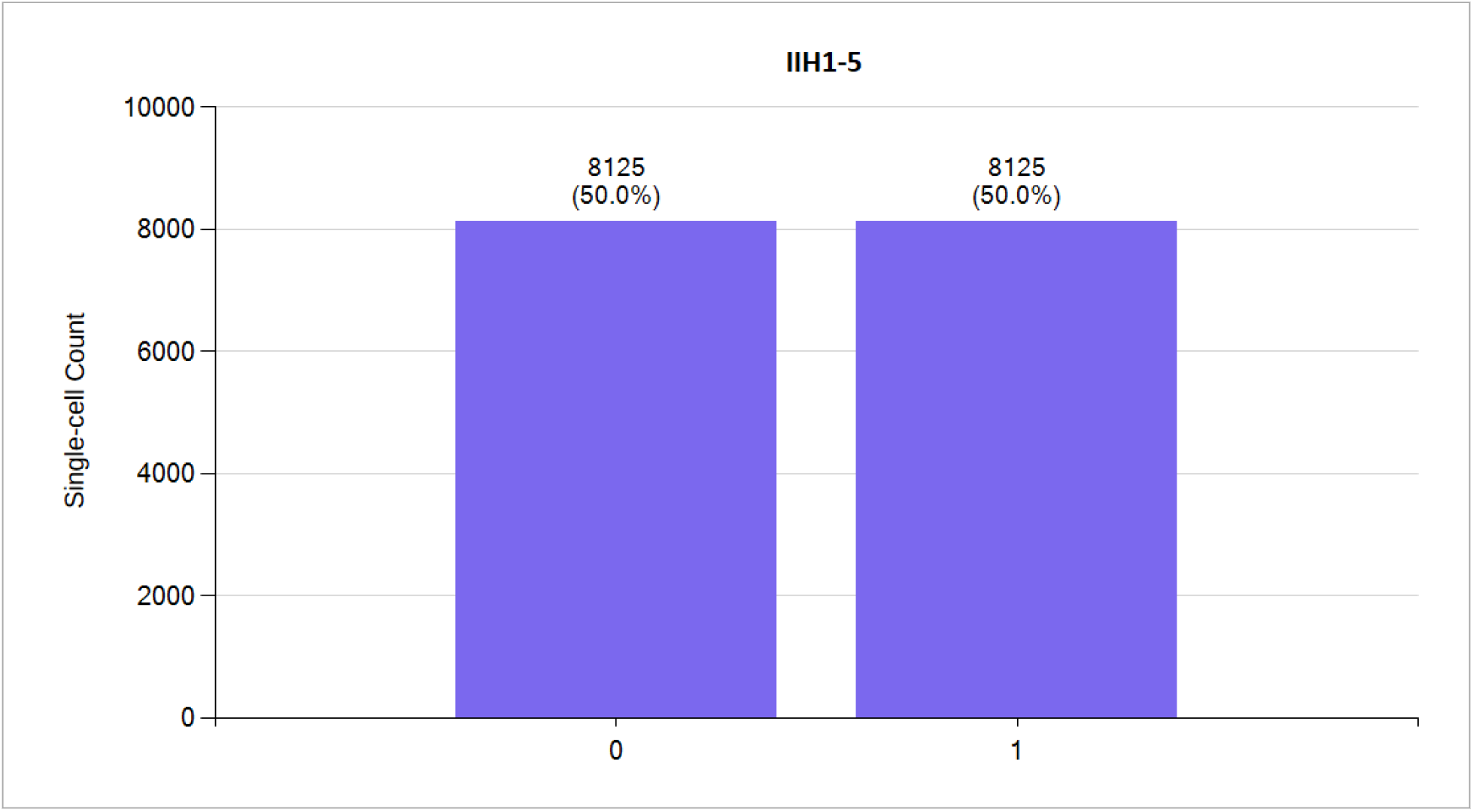
Blood leukocytes single-cell count for five IIH patients divided to two subsets randomly (0,1).

**Figure 6:**
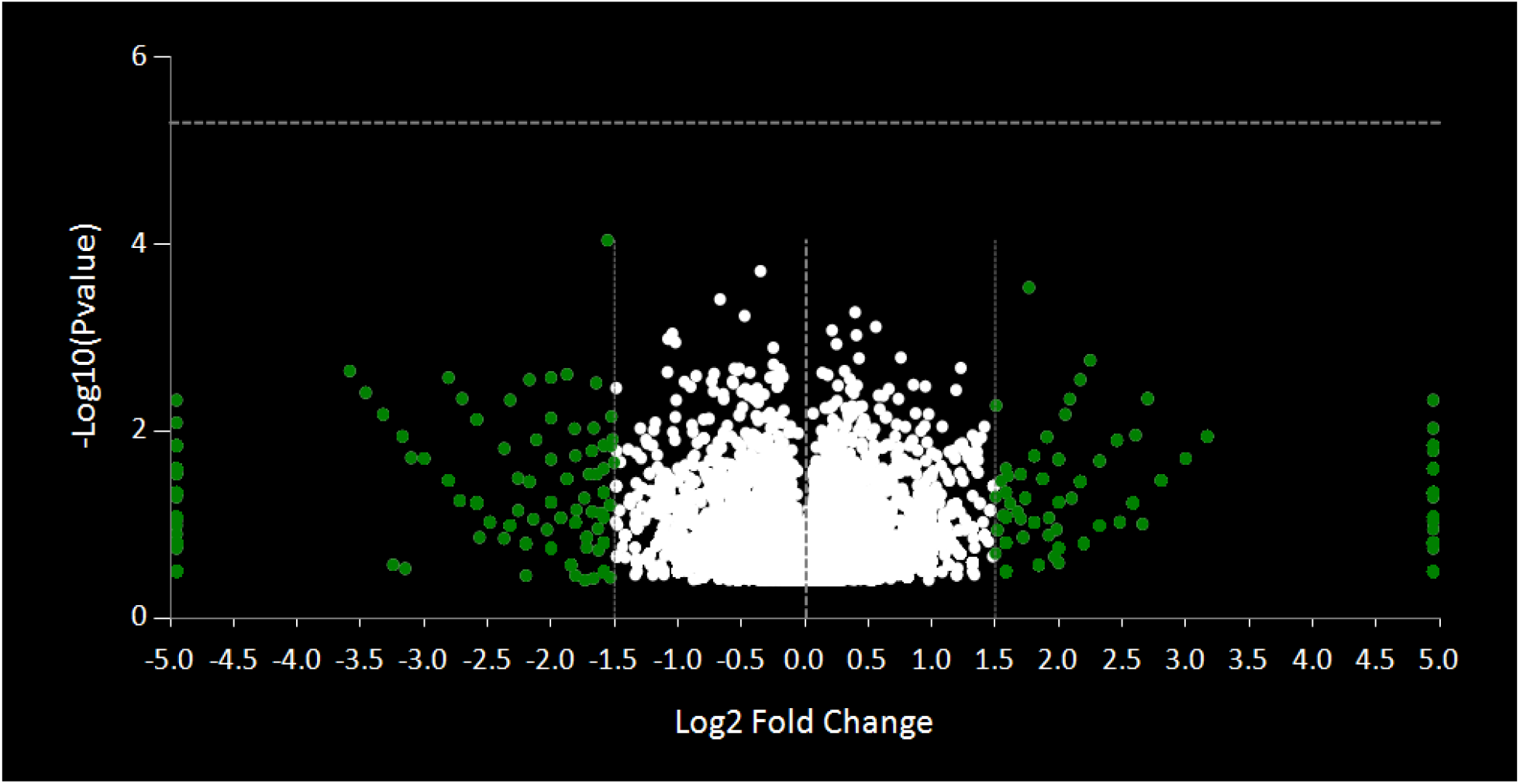
No significant upregulated or downregulated genes within the two randomly selected subsets of blood leukocytes from five IIH patients.

### Differential Gene Expression – Comparing Single Cells between MS and IIH patients

Furthermore, we conducted a comparative analysis of single cells obtained from IIH patients and MS patients. This investigation aimed to identify differentially expressed genes in both blood ( **Figure 7**) and CSF (**Figure 8**). This statistical comparison provides insight into molecular distinctions between blood and CSF single-cell leukocytes for improved understanding of MS pathophysiology.

**Figure 7:**
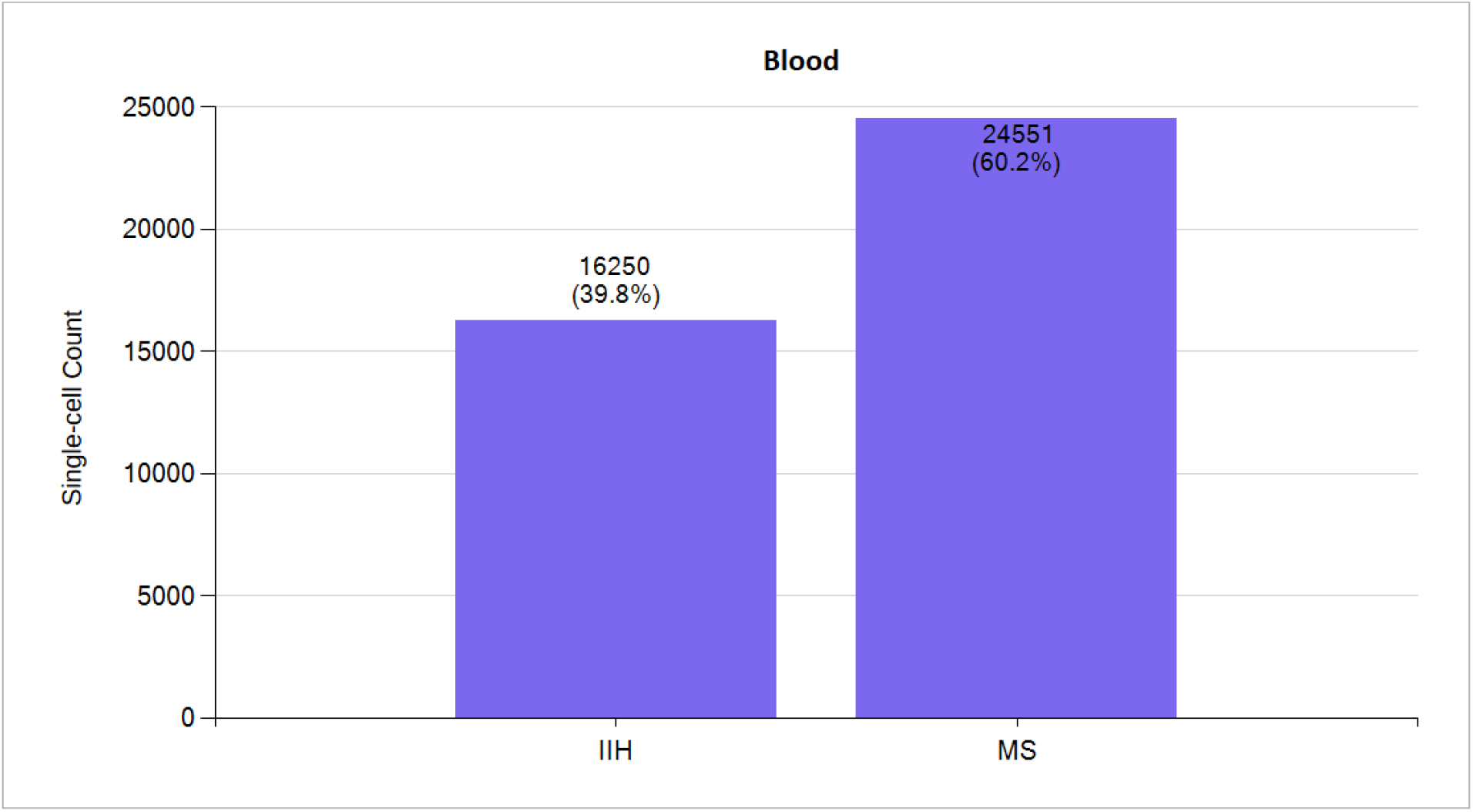
Single-cell count from blood leukocytes for five MS patients and five IIH patients.

**Figure 8:**
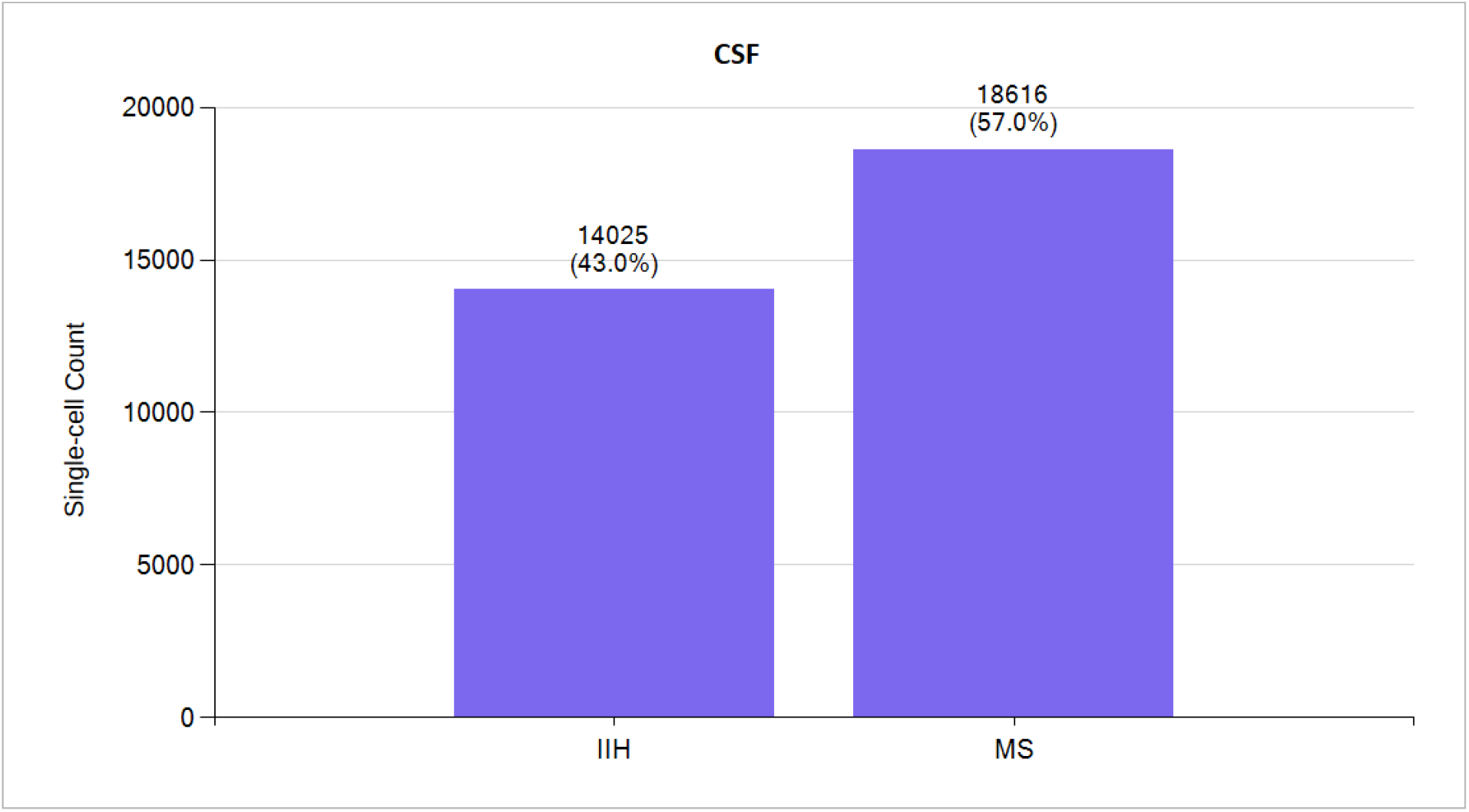
Single-cell count from CSF for five IIH patients and six MS patients.

**Table 1** presents the top 10 upregulated genes identified through comparative analysis of single-cell transcriptomes between MS and IIH patients in both blood and CSF samples. the upregulation of genes associated with the major histocompatibility complex (*HLA-C, HLA-B, HLA-A*) in MS compared to IIH suggests a potentially heightened immune activation or antigen presentation in MS [2]. Additionally, the differential expression of genes involved in ribosome biogenesis (*RPS26, NOP53*) and cellular proliferation regulation (*BTG1*) may reflect underlying differences in cellular turnover and growth processes between the two conditions [3]. Blood and CF samples shares three genes, *HLA-B, BTG1* and *HNRNPF*. Heterogenous nuclear ribonucleoproteins (*HNRNPs*) are a complex and functionally diverse family of RNA binding proteins with multifarious roles. They are involved, directly or indirectly, in alternative splicing, transcriptional and translational regulation, stress granule formation, cell cycle regulation, and axonal transport. It is unsurprising, given their heavy involvement in maintaining functional integrity of the cell, that their dysfunction has neurological implications [4].

**Table 1:** Top ten upregulated differentially expressed genes in MS patients compared to IIH patients.

| Top 10 Upregulated Genes in MS compared to IIH |  |  |  |
| --- | --- | --- | --- |
| Blood |  | CSF |  |
| Gene Symbol | Gene Name | Gene Symbol | Gene Name |
| RPS26 | Ribosomal Protein S26 | HLA-C | Major Histocompatibility Complex, Class I, C |
| GIMAP7 | GTPase, IMAP Family Member 7 | **HLA-B** | Major Histocompatibility Complex, Class I, B |
| GIMAP4 | GTPase, IMAP Family Member 4 | **BTG1** | BTG Anti-Proliferation Factor 1 |
| **BTG1** | BTG Anti-Proliferation Factor 1 | HLA-A | Major Histocompatibility Complex, Class I, A |
| HSPA8 | Heat Shock Protein Family A (Hsp70) Member 8 | IGKC | Immunoglobulin Kappa Constant |
| NOP53 | NOP53 Ribosome Biogenesis Factor | ACTB | Actin Beta |
| TMSB4X | Thymosin Beta 4 X-Linked | **HNRNPF** | Heterogeneous Nuclear Ribonucleoprotein F |
| **HLA-B** | Major Histocompatibility Complex, Class I, B | CMPK1 | Cytidine/Uridine Monophosphate Kinase 1 |
| **HNRNPF** | Heterogeneous Nuclear Ribonucleoprotein F | UBC | Ubiquitin C |
| ACTR3 | Actin Related Protein 3 | TRAC | T Cell Receptor Alpha Constant |

**Table 2** presents the top 10 downregulated genes in MS compare to IIH single cells (leukocytes), with a focus on blood and CSF samples. In blood, genes like HBB and HBA2 exhibit significant downregulation, suggesting potential alterations in oxygen transport and metabolism [5]. Similarly, in CSF, genes like *RPL37A* and *RPL38* show notable downregulation, indicating possible disruptions in ribosomal function and protein synthesis [3]. In addition to the hemoglobin subunits and ribosomal proteins mentioned, the downregulated genes in both blood and CSF samples encompass a variety of functional categories. For instance, *CD69* is involved in immune cell activation and differentiation, potentially indicating altered immune response dynamics in MS compared to IIH. *EIF5A* plays a crucial role in protein synthesis initiation, suggesting dysregulation in cellular translational processes. *TPT1* is associated with cell growth, differentiation, and apoptosis, implicating potential disruptions in cellular homeostasis. Additionally, *CXCR4* is a chemokine receptor involved in cell migration and inflammation, hinting at possible differences in inflammatory signaling pathways between MS and IIH. Furthermore, genes like *TSC22D3* and *CASK* may contribute to various cellular processes, including transcriptional regulation and synaptic function, highlighting the multifaceted nature of the molecular alterations observed in MS compared to IIH.

**Table 2:**
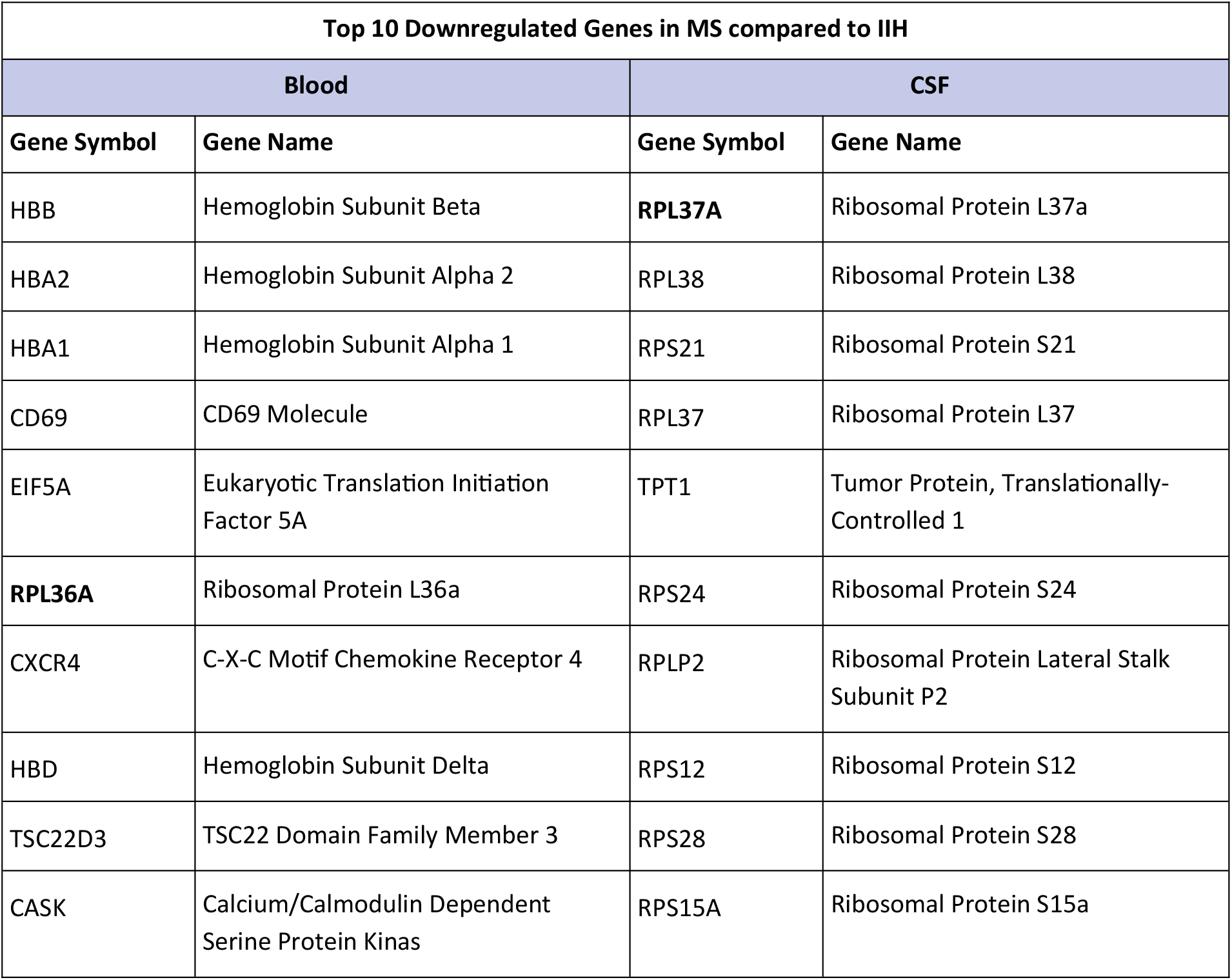
Top ten downregulated differentially expressed genes in MS patients compared to IIH patients.

| Top 10 Downregulated Genes in MS compared to IIH |  |  |  |
| --- | --- | --- | --- |
| Blood |  | CSF |  |
| Gene Symbol | Gene Name | Gene Symbol | Gene Name |
| HBB | Hemoglobin Subunit Beta | **RPL37A** | Ribosomal Protein L37a |
| HBA2 | Hemoglobin Subunit Alpha 2 | RPL38 | Ribosomal Protein L38 |
| HBA1 | Hemoglobin Subunit Alpha 1 | RPS21 | Ribosomal Protein S21 |
| CD69 | CD69 Molecule | RPL37 | Ribosomal Protein L37 |
| EIF5A | Eukaryotic Translation Initiation Factor 5A | TPT1 | Tumor Protein, Translationally-Controlled 1 |
| **RPL36A** | Ribosomal Protein L36a | RPS24 | Ribosomal Protein S24 |
| CXCR4 | C-X-C Motif Chemokine Receptor 4 | RPLP2 | Ribosomal Protein Lateral Stalk Subunit P2 |
| HBD | Hemoglobin Subunit Delta | RPS12 | Ribosomal Protein S12 |
| TSC22D3 | TSC22 Domain Family Member 3 | RPS28 | Ribosomal Protein S28 |
| CASK | Calcium/Calmodulin Dependent Serine Protein Kinas | RPS15A | Ribosomal Protein S15a |

**Table 3** outlines the top 10 upregulated gene ontology terms observed in MS compared to IIH single cells, with a focus on blood and CSF. In both blood and CSF, several biological processes (BP) related to cellular differentiation and apoptotic regulation stand out, including positive regulation of myoblast and endothelial cell differentiation, along with positive regulation of fibroblast apoptotic processes [6]. These findings suggest potential alterations in tissue remodeling and cellular survival mechanisms. Furthermore, molecular function (MF) terms such as TAP complex binding and TAP binding highlight potential changes in antigen processing and presentation pathways, which could influence immune responses [7]. Additionally, terms related to cytotoxicity, calcium ion regulation, and apoptotic signaling pathways point towards intricate molecular changes underlying the pathogenesis of MS, potentially providing valuable insights into its distinct clinical features. Moreover, cellular component (CC) terms like presynaptic cytosol indicate potential involvement of synaptic structures, suggesting neuronal alterations in MS [8]. Overall, these upregulated gene ontology terms offer a glimpse into the complex molecular landscape driving the pathophysiology of MS and signify promising avenues for further research into its underlying mechanisms.

**Table 3:** Top ten upregulated gene ontology terms in MS patients compared to IIH patients.

| Top 10 Upregulated Gene Ontology Terms in MS compared to IIH |  |
| --- | --- |
| Blood | CSF |
| (BP) positive regulation of myoblast differentiation | **(BP) positive regulation of fibroblast apoptotic process** |
| (BP) negative regulation of mitotic cell cycle | **(BP) positive regulation of endothelial cell differentiation** |
| **(BP) positive regulation of endothelial cell differentiation** | (MF) TAP complex binding |
| **(BP) positive regulation of fibroblast apoptotic process** | (BP) T cell mediated cytotoxicity directed against tumor cell target |
| (BP) release of sequestered calcium ion into cytosol | (BP) antigen processing and presentation of endogenous peptide antigen via MHC class I via ER pathway |
| (BP) regulation of aerobic respiration | (MF) TAP binding |
| (MF) C3HC4-type RING finger domain binding | (MF) CD8 receptor binding |
| (BP) positive regulation of protein K63-linked deubiquitination | (BP) regulation of intrinsic apoptotic signaling pathway in response to DNA damage by p53 class medi |
| (BP) negative regulation of signal transduction by p53 class mediator | (BP) regulation of interleukin-12 production |
| (CC) presynaptic cytosol | (BP) regulation of dendritic cell differentiation |

**Table 4** illustrates the top 10 downregulated gene ontology terms observed in IIH compared to MS single cells, focusing on blood and cerebrospinal fluid (CSF) samples. These findings unveil potential molecular alterations underlying the pathogenesis of IIH, shedding light on the nuanced differences between IIH and MS. Notably, biological processes (BP) related to various cellular functions, such as response to ultrasound [9], CXCL12-activated CXCR4 signaling pathway [10], and telencephalon cell migration, are markedly downregulated in both blood and CSF samples. These changes suggest potential disruptions in cellular signaling cascades, migration dynamics, and developmental processes in IIH. Moreover, terms related to ribosome assembly, including eukaryotic 80S initiation complex and 90S preribosome assembly, indicate potential dysregulation in protein synthesis machinery [11], which may impact cellular homeostasis and function. Additionally, terms associated with neuronal functions, such as neuron recognition and myelin maintenance [12], highlight potential alterations in neuronal integrity and myelination processes in IIH, which could contribute to its distinct clinical manifestations compared to MS. Furthermore, downregulation of terms related to immune regulation, such as negative regulation of activation-induced cell death of T cells, suggests potential immunomodulatory mechanisms underlying IIH pathology [13].

**Table 4:** Top ten downregulated gene ontology terms in MS patients compared to IIH patients.

| Top 10 Downregulated Gene Ontology terms in MS compared to IIH |  |
| --- | --- |
| Blood | CSF |
| (BP) response to ultrasound | (CC) eukaryotic 80S initiation complex |
| (BP) CXCL12-activated CXCR4 signaling pathway | (BP) 90S preribosome assembly |
| (MF) C-X-C motif chemokine 12 receptor activity | (BP) ribonucleoprotein complex assembly |
| (BP) telencephalon cell migration | (BP) axial mesoderm development |
| (BP) neuron recognition | (BP) negative regulation of intrinsic apoptotic signaling pathway in response to DNA damage |
| (BP) positive regulation of mesenchymal stem cell migration | (BP) endonucleolytic cleavage to generate mature 3-end of SSU-rRNA from (SSU-rRNA, 5.8S rRNA, LSU-rRNA |
| (MF) myosin light chain binding | (BP) middle ear morphogenesis |
| (BP) negative regulation of activation-induced cell death of T cells | (BP) endonucleolytic cleavage in ITS1 to separate SSU-rRNA from 5.8S rRNA and LSU-rRNA from transcript (SSU-rRNA, 5.8S rRNA, LSU-rRNA) |
| (BP) detection of temperature stimulus involved in sensory perception of pain | (BP) positive regulation of selenocysteine incorporation |
| (BP) myelin maintenance | (BP) ossification |

**Table 5** delineates the top 10 upregulated Reactome pathways observed in MS compared to IIH single cells, with a focus on blood and CSF samples. The upregulation of *FOXO-mediated transcription of cell cycle genes* shared in both blood and CSF leukocytes suggests potential alterations in cell cycle regulation [14]. Similarly, increased activity in *Neurotransmitter release cycle* and *GABA synthesis* pathways may reflect alterations in neuronal communication and neurotransmitter balance in MS [15]. Additionally, the upregulation of pathways related to *Antigen presentation, Interferon signaling*, and *Interleukin-2 signaling* underscores the involvement of immune regulatory mechanisms in the pathogenesis, hinting at potential autoimmune components in MS [16]. Moreover, *Lipophagy* and *Endosomal/vacuolar pathways* point towards potential disruptions in cellular metabolism and intracellular trafficking processes [17]. The heightened activity in pathways associated with *DNA damage response and senescence*, as well as regulation of *Heat shock response*, suggests potential cellular stress and adaptive responses [18].

**Table 5:**
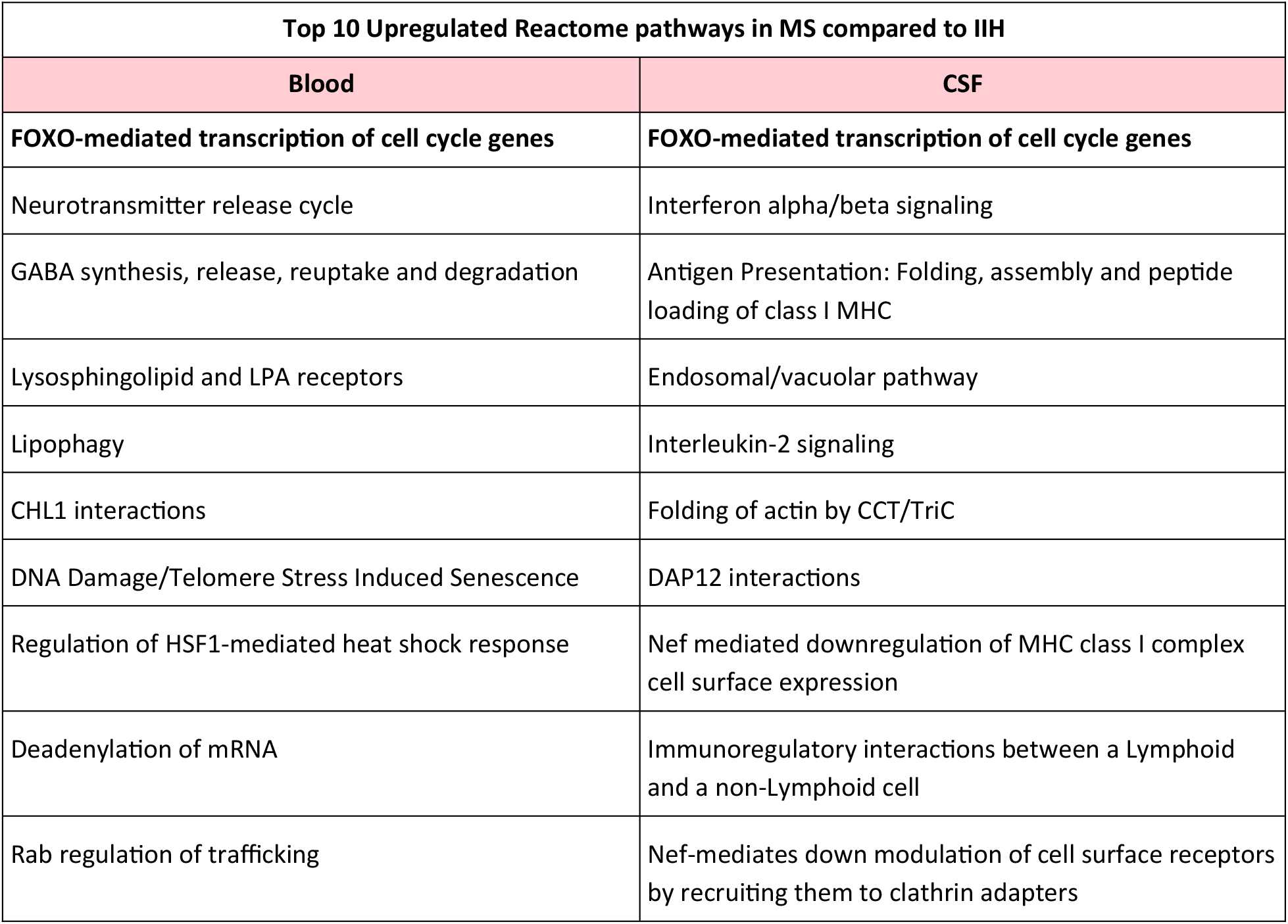
Top ten upregulated Reactome pathways in MS patients compared to IIH patients.

| Top 10 Upregulated Reactome pathways in MS compared to IIH |  |
| --- | --- |
| Blood | CSF |
| FOXO-mediated transcription of cell cycle genes | FOXO-mediated transcription of cell cycle genes |
| Neurotransmitter release cycle | Interferon alpha/beta signaling |
| GABA synthesis, release, reuptake and degradation | Antigen Presentation: Folding, assembly and peptide loading of class I MHC |
| Lysosphingolipid and LPA receptors | Endosomal/vacuolar pathway |
| Lipophagy | Interleukin-2 signaling |
| CHL1 interactions | Folding of actin by CCT/TriC |
| DNA Damage/Telomere Stress Induced Senescence | DAP12 interactions |
| Regulation of HSF1-mediated heat shock response | Nef mediated downregulation of MHC class I complex cell surface expression |
| Deadenylation of mRNA | Immunoregulatory interactions between a Lymphoid and a non-Lymphoid cell |
| Rab regulation of trafficking | Nef-mediates down modulation of cell surface receptors by recruiting them to clathrin adapters |

**Table 6** outlines the top 10 downregulated Reactome pathways observed in MS compared to IIH patients, using both blood and CSF single-cell leukocytes samples. These findings provide valuable insights into the molecular alterations underlying the pathogenesis of MS, highlighting distinct biological processes affected in MS compared to IIH. Notably, pathways related to erythrocyte function, such as E*rythrocytes take up oxygen and release carbon dioxide* and E*rythrocytes take up carbon dioxide and release oxygen* are downregulated in both blood and CSF samples, suggesting potential disruptions in oxygen transport dynamics in MS [19]. Additionally, pathways associated with mitotic spindle formation and chromosomal cohesion, such as *Amplification of signal from the kinetochores* and *Resolution of Sister Chromatid Cohesion* indicate potential dysregulation in cell division processes in MS, which could impact cellular proliferation and tissue homeostasis [20]. Furthermore, downregulation of pathways related to scavenger receptor function and small molecule transport suggests potential alterations in cellular metabolism and nutrient uptake mechanisms in MS. Interestingly, pathways associated with *Autophagy* and *Selenoamino acid metabolism*, such as *Chaperone Mediated Autophagy* and *Selenocysteine synthesis* are also downregulated, implying potential disruptions in cellular clearance mechanisms and antioxidant defenses in MS [21].

**Table 6:** Top ten downregulated Reactome pathways in MS patients compared to IIH patients.

| Top 10 Downregulated Reactome pathways in MS compared to IIH |  |
| --- | --- |
| Blood | CSF |
| Erythrocytes take up oxygen and release carbon dioxide | Amplification of signal from the kinetochores |
| Erythrocytes take up carbon dioxide and release oxygen | Amplification of signal from unattached kinetochores via a MAD2 inhibitory signal |
| O<sub>2</sub>/CO<sub>2</sub> exchange in erythrocytes | EML4 and NUDC in mitotic spindle formation |
| Binding and Uptake of Ligands by Scavenger Receptors | Mitotic spindle checkpoint |
| Scavenging of heme from plasma | Resolution of sister chromatid cohesion |
| Late endosomal microautophagy | Selenocysteine synthesis |
| Transport of small molecules | Viral mRNA translation |
| Factors involved in megakaryocyte development and platelet production | Selenoamino acid metabolism |
| Chaperone mediated autophagy | Eukaryotic translation termination |
| Autophagy | Response of EIF2AK4 (GCN2) to amino acid deficiency |

Overall, Reactome pathways provide important insights into the complex molecular landscape underlying MS pathology and offer potential targets for further investigation and therapeutic intervention.

### *CD69* and *HNRNPK* – Two potential Driver Genes

Finally, we conducted a thorough differentially expressed gene analysis, comparing one IIH patient against one MS patient, examining both blood and CSF samples. From this analysis, we pinpointed and selected the top 800 genes showing significant upregulation or downregulation in each comparison. Interestingly across all 25 comparisons (5 IIH patients and 5 MS patients) for the blood group only one gene *CD69* (downregulated in MS) remained consistently present, while in the CSF group, the gene *HNRNPK* (upregulated in MS) emerged as the sole common denominator across all 30 comparisons (5 IIH patients and 6 MS patients).

In MS brains, *CD69+* cells were detected almost exclusively in reactive normal appearing white matter, active and chronic active white matter lesions [22]. It has been shown that *CD69* knockout mice exhibit an enhanced susceptibility to different inflammatory diseases, mainly those mediated by Th17 lymphocytes. This underscores the intricate immune-regulatory function of *CD69* in humans, suggesting its potential as a target molecule for the treatment of immune-mediated disorders [23].

The *HNRNPK* gene is involved in multiple processes of gene expression, including chromatin remodeling and transcription. It also regulates inflammatory gene expression by mediating splicing patterns of transcriptional factors. Research indicates that dysfunction of a specific member within the heterogeneous nuclear ribonucleoproteins family (*HNRNPA1*), alters RNA splicing and drives neurodegeneration in MS [24].

## Discussion

The examination of single-cell transcriptomes data from patients diagnosed with MS and IIH illuminates the molecular landscapes characterizing these neurological conditions through differential gene expression analysis and exploration of gene ontology terms and Reactome pathways. The absence of significant differential gene expression within individual patients, within MS patients and within IIH patients, enhances the credibility of our single-cell data, underscoring the reliability of our findings. Furthermore, the comparative analysis between MS and IIH revealed distinct gene-expression profiles in both blood and CSF samples, highlighting molecular differences underlying the two conditions.

Our analysis identified upregulated genes associated with the major histocompatibility complex (MHC) in MS compared to IIH, suggesting heightened immune activation or antigen presentation in MS. Additionally, differential expression of genes involved in ribosome biogenesis and cellular proliferation regulation may reflect underlying differences in cellular turnover and growth processes between MS and IIH. Notably, the upregulation of heterogenous nuclear ribonucleoproteins (*HNRNPs*) in both blood and CSF samples underscores their multifaceted roles in cellular homeostasis and implicates their dysfunction in neurological implications, potentially contributing to MS pathology. Conversely, downregulated genes in MS compared to IIH encompass diverse functional categories, including those involved in oxygen transport, ribosomal function, immune cell activation, protein synthesis, cellular growth, differentiation, apoptosis, inflammation, and synaptic function.

Our gene ontology analysis revealed upregulated terms associated with cellular differentiation, apoptotic regulation, antigen processing, presentation pathways, cytotoxicity, calcium ion regulation, apoptotic signaling pathways, and synaptic structures in MS compared to IIH. These findings provide insights into the molecular landscape driving MS pathophysiology, suggesting alterations in tissue remodeling, cellular survival mechanisms, immune responses, inflammatory signaling pathways, and neuronal integrity. Additionally, downregulated gene ontology terms in MS compared to IIH shed light on potential disruptions in cellular signaling cascades, migration dynamics, developmental processes, protein synthesis machinery, neuronal integrity, myelination processes, and immune regulation underlying IIH pathology.

Additionally, analysis of Reactome pathways identified upregulated pathways associated with cell cycle regulation, neurotransmitter balance, antigen presentation, immune regulatory mechanisms, cellular metabolism, intracellular trafficking processes, DNA damage response, cellular stress, and adaptive responses in MS. Alternatively, downregulated Reactome pathways in MS compared to IIH indicate potential disruptions in oxygen transport dynamics, cell division processes, cellular metabolism, nutrient uptake mechanisms, cellular clearance mechanisms, and antioxidant defenses in MS. These findings underscore the complex molecular alterations underlying MS pathogenesis and highlight potential targets for therapeutic intervention.

Finally, in a comprehensive analysis, we compared gene expression profiles in blood and CSF samples from one IIH patient and one MS patient, identifying the top 800 differentially expressed genes in each comparison. Notably, *CD69* emerged as the sole consistently downregulated gene in MS across blood samples, while *HNRNPK* stood out as the consistently upregulated gene in MS across CSF samples. Studies on *CD69* knockout mice suggest its crucial role in regulating inflammatory diseases, particularly those mediated by Th17 lymphocytes, highlighting its potential as a therapeutic target for immune-mediated disorders. Additionally, *HNRNPK*, involved in various gene expression processes, including chromatin remodeling and transcription, appears to influence inflammatory gene expression in MS.

## Data Availability

All data produced are available online at:
https://www.ncbi.nlm.nih.gov/geo/query/acc.cgi?acc=GSE138266

## Summary

Our study provides a comprehensive analysis of single-cell transcriptomes data, elucidating gene-expression signatures, gene ontology terms, and Reactome pathways specific to MS pathophysiology.

Interestingly, we discovered *CD69* and *HNRNPK* as two potential driver genes in MS. By delineating molecular distinctions between MS and IIH, our findings advance our understanding of MS pathogenesis and offer potential targets for diagnostic and therapeutic strategies. Further research is warranted to validate these findings and explore their clinical implications in MS management.

## Notes

### Competing Interest Statement

The authors have declared no competing interest.

### Funding Statement

This study did not receive any funding

